# Knee-extensor strength, symptoms, and need for surgery after two, four, or six exercise sessions/week using a home-based *one*-exercise program: A randomized dose-response trial of knee-extensor resistance exercise in patients eligible for knee replacement (the QUADX-1 trial)

**DOI:** 10.1101/2021.04.07.21254965

**Authors:** Rasmus Skov Husted, Anders Troelsen, Henrik Husted, Birk Mygind Grønfeldt, Kristian Thorborg, Thomas Kallemose, Michael Skovdal Rathleff, Thomas Bandholm

**Affiliations:** Department of Clinical Research, Copenhagen University Hospital Amager-Hvidovre, Hvidovre, Denmark; Physical Medicine & Rehabilitation Research – Copenhagen (PMR-C); Department of Physical and Occupational Therapy; Department of Clinical Research; Department of Orthopedic Surgery, Copenhagen University Hospital Amager-Hvidovre, Hvidovre, Denmark; Clinical Orthopedic Research Hvidovre (CORH), Department of Orthopedic Surgery, Copenhagen University Hospital Amager-Hvidovre, Hvidovre, Denmark; Sports Orthopaedic Research Center – Copenhagen (SORC-C), Department of Orthopedic Surgery, Copenhagen University Hospital Amager-Hvidovre, Hvidovre, Denmark; Center for General Practice at Aalborg University, Aalborg, Denmark; Department of Occupational Therapy and Physiotherapy, Aalborg University Hospital, Aalborg, Denmark; Department of Health Science and Technology, Aalborg University, Denmark; Department of Clinical Medicine, University of Copenhagen, Denmark

**Author notes:** E-mail addresses: Anders Troelsen, Henrik Husted, Birk Mygind Grønfeldt, Kristian Thorborg, Thomas Kallemose, Michael Skovdal Rathleff, Thomas Bandholm. Primary investigator and corresponding author. Department of Clinical Research (Section 056), Copenhagen University Hospital Amager-Hvidovre, Kettegaard Allé 30, 2650 Hvidovre, Denmark. Mail.

**Keywords:** Knee osteoarthritis, knee-extensor resistance exercise, dose-response, knee replacement, coordinated non-surgical and surgical care

## Abstract

**Objective:** To investigate firstly the efficacy of three different dosages of *one* home-based, knee-extensor resistance exercise on knee-extensor strength in patients eligible for knee replacement, and secondly, the influence of exercise on symptoms, physical function and decision on surgery.

**Method:** One-hundred and forty patients eligible for knee replacement were randomized to three groups: 2, 4 or 6 home-based knee-extensor resistance exercise-sessions per week (group 2, 4 and 6 respectively) for 12 weeks. Primary outcome: isometric knee-extensor strength. Secondary outcomes: Oxford Knee Score, Knee injury and Osteoarthritis Outcome Score, average knee pain last week (0-10 numeric rating scale), 6-minute walk test, stair climbing test, exercise adherence and “need for surgery”.

**Results:** Primary analysis: Intention-to-treat analysis of 140 patients did not find statistically significant differences between the groups from baseline to after 12 weeks of exercise in isometric knee-extensor strength: Group 2 vs. 4 (0.003 Nm/kg (0.2%) [95% CI -0.15 to 0.15], P=0.965) and group 4 vs. 6 (−0.04 Nm/kg (−2.7%) [95% CI -0.15 to 0.12], P=0.628). Secondary analysis: Intention-to-treat analyses showed statistically significant differences between the two and six sessions/week groups in favor of the two sessions/week group for Oxford Knee Score: 4.8 OKS points (15.2%) [1.3 to 8.3], P=0.008) and avg. knee pain last week (NRS 0-10): - 1.3 NRS points (−19.5%) [-2.3 to -0.2], P=0.018. After the 12-week exercise intervention, data were available for 117 patients (N=39/group): 38 (32.5%) patients wanted surgery and 79 (67.5%) postponed surgery. This was independent of exercise dosage.

**Conclusion:** In patients eligible for knee-replacement we found no between-group differences in isometric knee extensor strength after 2, 4 and 6 knee-extensor resistance exercise sessions per week. We saw no indication of an exercise dose-response relationship for isometric knee-extensor strength and only clinically irrelevant within group changes. For some secondary outcome (e.g. KOOS subscales) we found clinically relevant within group changes, which could help explain why only one in three patients decided to have surgery after the simple home-based exercise intervention.

**Trial registration:** ClinicalTrials.gov identifier: NCT02931058. Preprint: https://doi.org/10.1101/2021.04.07.21254965.

## Introduction

Exercise therapy can reduce symptoms and postpone surgery in about 50% of patients with knee OA^1–4^ and guidelines recommend that exercise therapy is tried out before surgery is considered in patients eligible for knee replacement.^5–10^ Because the indication for knee replacement is not clear-cut, identifying the right patients to operate at the right time is difficult^11,12^ – making the coordination of non-surgical and surgical care crucial in selecting the right candidates for knee replacement.^13–16^ Any changes in symptoms after exercise therapy may play an important role in the shared decision-making process for surgery.^17–20^

Exercise programs for patients with knee OA like ‘Good Life with osteoArthritis in Denmark’ (GLA:D) - successfully implemented worldwide^21^ - and ‘Better management of patients with OsteoArthritis’ (BOA) support the effectiveness of exercise therapy and education for these patients and deliver optimized care.^22–25^ The exercise programs are supervised, require physical attendance at fixed times and often require self-payment; factors which can be barriers for some patients and hinder participation and long-term adherence, creating inequality for the care accessible.^26–29^ An important element in exercise programs for patients with knee OA is knee-extensor strength^30^, as decreased knee-extensor strength is associated with an increased risk of developing knee OA,^31^ risk of knee pain and decline in function.^32^ According to the American College of Sports Medicine (ACSM) two exercise sessions per week is the recommended minimum dosage required for muscle strength gains, four is likely optimal, and six is likely to have no additional benefit, but could increase pain.^33,34^ Based on this, we investigated the dose-response relationship of *one* home-based resistance exercise targeting the knee-extensor muscles, using a very simple and low-cost exercise option. Compared to supervised exercise programs, this solution does not require physical attendance at fixed times and is free of charge – providing patients with an alternative treatment option.

We asked the following:

1. Is there a dose-response relationship between knee-extensor resistance exercise and change in isometric knee-extensor strength in patients eligible for knee replacement?
2. Do different dosages of simple knee-extensor resistance exercise change symptoms and decision on surgery in patients eligible for knee replacement?

The primary aim was to investigate the efficacy of three different dosages of home-based, knee-extensor resistance exercise on isometric knee-extensor strength in patients eligible for knee replacement due to severe knee OA, and secondly, to investigate the influence of exercise on symptoms, physical function and decision on surgery. The hypothesis was that an exercise dosage of four knee-extensor resistance exercise sessions per week would elicit the greatest change in isometric knee-extensor strength pre-operatively compared to two or six sessions per week.

## Methods

### Trial design

The QUADX-1 trial is a three-arm parallel-group randomized dose-response trial with three intervention groups and no control group. The trial was pre-registered on clinicaltrials.gov on 10^th^ October, 2016 (NCT02931058) before enrollment of the first patient, and the full trial protocol – including protocol amendments – was published 18^th^ January, 2018.^35^ Approvals from the Ethics Committee of the Capital Region, Denmark (H-16025136) and the Danish Data Protection Agency (2012-58-0004) were obtained before the first patient was enrolled.

### Trial amendments

Due to an oversight, the second research question and purpose were not pre-registered. Hence, we consider them secondary and exploratory. All other trial amendments are reported in the trial protocol.^35^

### Participants

Patients potentially eligible for trial participation were recruited at the surgical outpatient clinic. The inclusion criteria were: eligible for knee replacement due to knee OA (assessed by an orthopedic surgeon), radiographically verified knee OA with Kellgren-Lawrence classification ≥ 2 (Kellgren-Lawrence scores 2 were included to mimic everyday clinical practice),^36,37^ average knee pain ≥ 3 (Numeric Rating Scale (NRS)) in the last week, eligible for home-based knee-extensor resistance exercise, age ≥ 45 years, resident in one of three municipalities involved in the trial (Copenhagen, Hvidovre or Broendby) and able to speak and understand Danish. The exclusion criteria were: exercise therapy being contra-indicated, neurological disorder, diagnosed systemic disease (American Society of Anesthesiologists’ physical status classification score ≥ 4),^38^ terminal illness, severe bone deformity demanding use of non-standard implants, or a greater weekly alcohol consumption than the national recommendation.^39^

### Interventions

Following baseline assessment, the patients were referred to a physiotherapist in their local municipal rehabilitation setting. Here the patients were instructed how to perform a single knee-extensor resistance exercise at home. The knee-extensor resistance exercise was performed sitting on a chair with an exercise band wrapped around the ankle and fixed behind a door for resistance. Patients were provided with a personal exercise band for exercising at home and a brochure with instructional notes and illustrations. The patients were randomized to one of three exercise dosage groups for twelve weeks: the two sessions/week group, the four sessions/week group or the six sessions/week group. For all groups, training comprised only the single knee-extensor resistance exercise. Patients were instructed to perform the exercise in three sets of twelve repetitions with each repetition lasting eight seconds (concentric phase 3 s, isometric phase 1 s, eccentric phase 4 s). The intervention was personalized to the extent where each patient was exercising with an individual absolute resistance corresponding to a relative load of twelve repetition maximum (RM). The patients were instructed to continue until volitional muscular failure. That is, until the knee-extensor muscles were maximally fatigued, and they were not able to perform further repetitions. If volitional muscular failure occurred before twelve RM, the resistance of the elastic band was adjusted so that the pre-determined number of repetitions could be completed (decrease in distance between the two endpoints of the elastic band). Whenever the resistance in the elastic band became too low (i.e., more than twelve repetitions per set could be performed), the patients were instructed to increase the resistance in the elastic band to achieve a new resistance corresponding to a relative load of twelve RM (increase in distance between the two endpoints of the elastic band). Detailed intervention description can be found in the trial protocol^35^ and a walkthrough video of the exercise is freely available online (https://bit.ly/3i59CJn).

### Assessments and outcomes

Outcomes were assessed: at baseline (t_0_), after twelve weeks of home-based exercise/before surgery (t_1_), at hospital discharge (1-8 days after surgery) (t_2_) and three months after surgery (t_3_). Outcomes at endpoints t_2_ and t_3_ were only collected for patients that underwent surgery. The primary endpoint was after the exercise period (t_1_) and the secondary endpoints were just before hospital discharge (t_2_) and three months after surgery (t_3_). After the 12-week exercise period, at endpoint t_1_, each patient’s decision on surgery was re-evaluated in a shared decision-making process between the patient and orthopedic surgeon (i.e. continue with exercise therapy or schedule knee replacement). Outcome assessments were performed blinded by the primary investigator and a research assistant dedicated to the trial.

### Primary outcome

The primary outcome was change in isometric knee-extensor strength from baseline to after the exercise period (t_0_-t_1_). Isometric knee-extensor strength was measured using a computerized strength chair (Good Strength Chair, Metitur Oy, Jyvaskyla, Finland), which is valid and reliable in the knee replacement population.^40^ Five measurements of maximal isometric knee-extensor strength at 60° knee flexion were completed, separated by 60-s pauses. The patients were instructed to extend their knee as forcefully as possible with a gradual increase in force over a 5-s period while receiving strong standardized verbal encouragement. Isometric knee-extensor strength is expressed as the maximal voluntary torque per kilogram body mass (Nm/kg). The highest obtained value was used for analysis.

### Secondary outcomes

The secondary outcomes were change in performance-based function comprising six-minute walk test (6MWT) and stair climb test (SCT), self-reported disability; Knee injury and Osteoarthritis Outcome Score (KOOS), Oxford Knee Score (OKS), current knee pain and average knee pain during the last week (0-10 NRS), “need for surgery” and objectively measured exercise adherence (t_0_-t_1_, t_0_-t_2_ and t_0_-t_3_). Other outcomes were registration of adverse events and harms.

The “need for surgery” outcome was an assessment of the patients’ self-perceived need for surgery. After the 12-week exercise period at outcome assessment t_1_ the patients were asked by the outcome assessor: *“Based on your knee symptoms in the last week would you say that you need knee surgery?”* Three answer options were possible: 1) *Yes, I believe I need surgery*, 2) *I do not know* or 3) *No, I do not believe I need surgery*.

Exercise therapy adherence was objectively quantified using a sensor attached to the exercise band (BandCizer© sensor technology).^41–43^ The sensor collects and stores data on date, time, number of sets, repetitions and time-under-tension (TUT). Patients were defined as adherent if >75% of the prescribed exercise sessions were completed.

Detailed information on the secondary outcomes is reported in the protocol paper.^35^

### Sample size

The sample size was calculated for a test of superiority (four knee-extensor resistance exercise sessions per week is superior to two or six sessions per week). For the primary planned three-group one-way ANOVA analysis, a sample size of 126 patients (42 per group) was required to obtain a power of 80%. The a priori sample size calculation was based on a normal mean difference with a two-sided significance level of 0.025 (Bonferroni correction for two tests (2 vs. 4 and 4 vs. 6), a minimal clinical important difference (MCID) of 0.15 Nm/kg (15%) and a common standard deviation of 0.22 Nm/kg in isometric knee-extensor strength.^44^ To allow for a dropout rate of 10%, a total of 140 patients were included in the intention-to-treat (ITT) analysis.

### Randomization

The patients were randomly assigned by a 1:1:1 allocation ratio. The random allocation sequence was computer-generated using simple (unrestricted) randomization by a statistician otherwise not involved in the trial. One hundred and forty sequentially numbered sealed opaque envelopes were generated. When a patient was included in the trial a research assistant independent of the trial opened an envelope and informed the patient’s municipality of the exercise group allocation.

### Blinding

All outcome assessors and the data analysts were blinded to the exercise group allocation. At outcome assessments the assessors started by informing the patients not to mention their exercise dosage. For analysis, the data was coded to conceal group allocation, blinding the data assessors and analysts to the patients’ allocation. The physiotherapists and patients were not blinded to the allocation due to the nature of the intervention, however, the patients were blinded to the other exercise dosages and the study hypothesis.

### Statistics

The primary intention-to-treat superiority analysis tested the hypothesis that an exercise dosage of four knee-extensor resistance exercise sessions per week would elicit a greater change in isometric knee-extensor strength pre-operatively compared to two or six sessions per week. For all outcomes, between group contrasts were compared using analysis of variance (one-way ANOVA). Normality assumptions of the model residuals were checked to ensure that the underlying assumptions of the statistical model were met. Normal distribution of data was checked by q-q plots and histograms. Analyses were adjusted for the following baseline variables: isometric knee-extensor strength, KOOS symptoms, KOOS ADL, KOOS sport and 6MWT. These adjustments were not prespecified. In a secondary analysis, the two sessions/week and six sessions/week groups were compared and follow the same principles as the primary analysis. As supplementary analyses, simple regression models were performed using the pooled exercise adherence data across all three groups. The dependent variables were the primary and secondary outcomes and the independent variable was exercise adherence quantified in two ways: 1) as total number of completed exercise sessions and 2) as total time-under-tension (TUT) per patient. All analyses followed the ITT principle and to create full datasets, missing data were imputed using multiple imputation (100 imputation sets). Multiple imputation models were based on age, gender, group allocation and all previous scores in relevant outcomes. Missing data break down is presented in Supplement 1. All analyses followed the pre-specified analysis plan^35^ and were performed in SAS Enterprise Guide 7.1.

## Results

### Participants

Between 25^th^ October 2016 and 8^th^ January 2019, 898 patients potentially eligible for knee replacement were assessed for eligibility. One-hundred and forty patients were included and randomized (Figure 1). Assessments at the primary endpoint (after 12 weeks of exercise [t_1_]) was completed for 117 patients (39/group). At the two secondary endpoints, 32 patients were available for assessment. Reasons for drop-out and missing data are provided in figure 1. Baseline characteristics are provided in table 1 and in Supplement 2.

**Figure 1.**
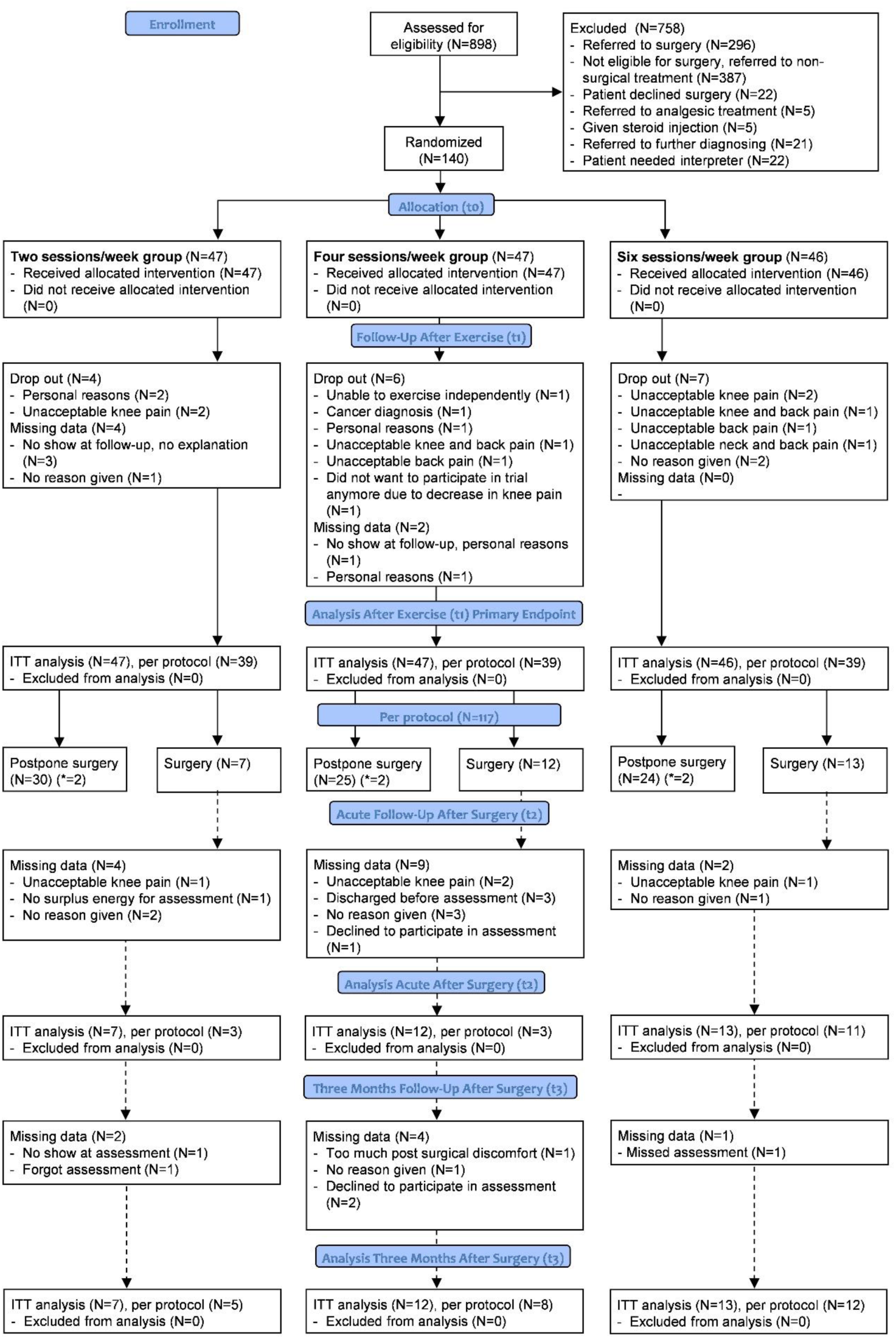
Flow chart of each assessment time-point of the trial according to the CONSORT guidelines.^86^ ITT = intention-to-treat analysis. Dotted lines indicate assessment time-points after surgery. *6 patients (N=2/group) wanted surgery but had competing co-morbidities disqualifying them as candidates for surgery (Supplement 6).

**Table 1.** Baseline characteristics (t_0_).

| Characteristics | All patients<br>N = 140 | Two sessions/week N<br>= 47 | Four sessions/week N<br>= 47 | Six sessions/week<br>N = 46 |
| --- | --- | --- | --- | --- |
| Gender (f/m) | 76 (54)/64 (46) | 22 (47)/25 (53) | 29 (62)/18 (38) | 25 (54)/21 (46) |
| Age (years) | 66.7 (9.9) | 67.5 (9.7) | 66.8 (10.0) | 65.8 (10.0) |
| Weight (kg) | 91.9 (19.9) | 92.1 (17.0) | 94.2 (21.8) | 89.8 (20.3) |
| Height (cm) | 169.2 (8.3) | 168.7 (7.0) | 170.1 (7.7) | 169.1 (9.9) |
| Municipality (Cph/Hvi/Bro) | 74 (53)/44<br>(31)/22 (16) | 22 (47)/16 (34)/9 (19) | 23 (49)/16 (34)/8 (17) | 29 (63)/12 (26)/5<br>(11) |
| Kellgren and Lawrence score<br>(2/3/4)* | 20 (15)/61<br>(44)/57 (41) | 5 (11)/20 (44)/21 (45) | 9 (20)/19 (41)/18 (39) | 6 (13)/22 (48)/18<br>(39) |
| Current knee pain (NRS 0-10) | 2.2 (2.2) | 2.1 (2.4) | 2.1 (2.2) | 2.4 (2.1) |
| Avg. knee pain last week (NRS<br>0-10) | 5.8 (1.6) | 5.7 (1.6) | 5.8 (1.6) | 5.8 (1.4) |
| Isometric knee-extensor<br>strength (Nm/kg) | 1.27 (0.52) | 1.31 (0.57) | 1.22 (0.49) | 1.28 (0.51) |
| KOOS Symp (0-100) | 55.0 (18.8) | 58.9 (19.4) | 53.4 (16.7) | 52.9 (19.6) |
| KOOS Pain (0-100) | 49.7 (16.4) | 51.7 (16.5) | 48.2 (16.7) | 49.6 (15.5) |
| KOOS ADL (0-100) | 55.3 (17.5) | 57.7 (17.0) | 51.7 (17.5) | 56.3 (17.3) |
| KOOS Sport (0-100) | 21.0 (20.8) | 24.5 (23.6) | 16.8 (16.7) | 21.3 (20.1) |
| KOOS QoL (0-100) | 32.7 (16.3) | 35.4 (16.3) | 31.2 (16.1) | 31.1 (15.9) |
| OKS (0-48) | 24.8 (7.6) | 26.2 (7.3) | 23.2 (8.0) | 24.9 (7.0) |
| 6MWT (m) | 402.3 (105.3) | 416.5 (94.1) | 387.7 (112.2) | 402.1 (102.8) |
| SCT up (secs) | 9.4 (5.1) | 8.7 (5.1) | 10.3 (5.4) | 9.0 (4.6) |
| SCT down (secs) | 10.4 (6.7) | 8.9 (5.3) | 11.9 (7.9) | 10.4 (6.4) |

For continuous data mean and SD are provided. For categorical data N and % are provided. Abbreviations: Cph = Copenhagen, Hvi = Hvidovre, Bro = Broendby, NRS = Numeric Rating Scale, KOOS = Knee injury and Osteoarthritis Outcome Score, OKS = Oxford Knee Score, 6MWT = six-minute walk test, SCT = Stair climb test. \* = missing data on two patients.

### Assessment after exercise

Primary outcome: Intention-to-treat analysis did not find statistically significant differences between the groups in change between baseline and following 12 weeks of exercise (primary endpoint (t_0_-t_1_)) in *isometric knee-extensor strength*: two sessions/week group vs. four sessions/week group; 0.003 Nm/kg (0.2%) [95% CI -0.15 to 0.15], P=0.965, and four sessions/week group vs. six sessions/week group; -0.04 Nm/kg (−2.7%) [95% CI -0.15 to 0.12], P=0.628 (Figure 2) (Table 2).

**Figure 2.**
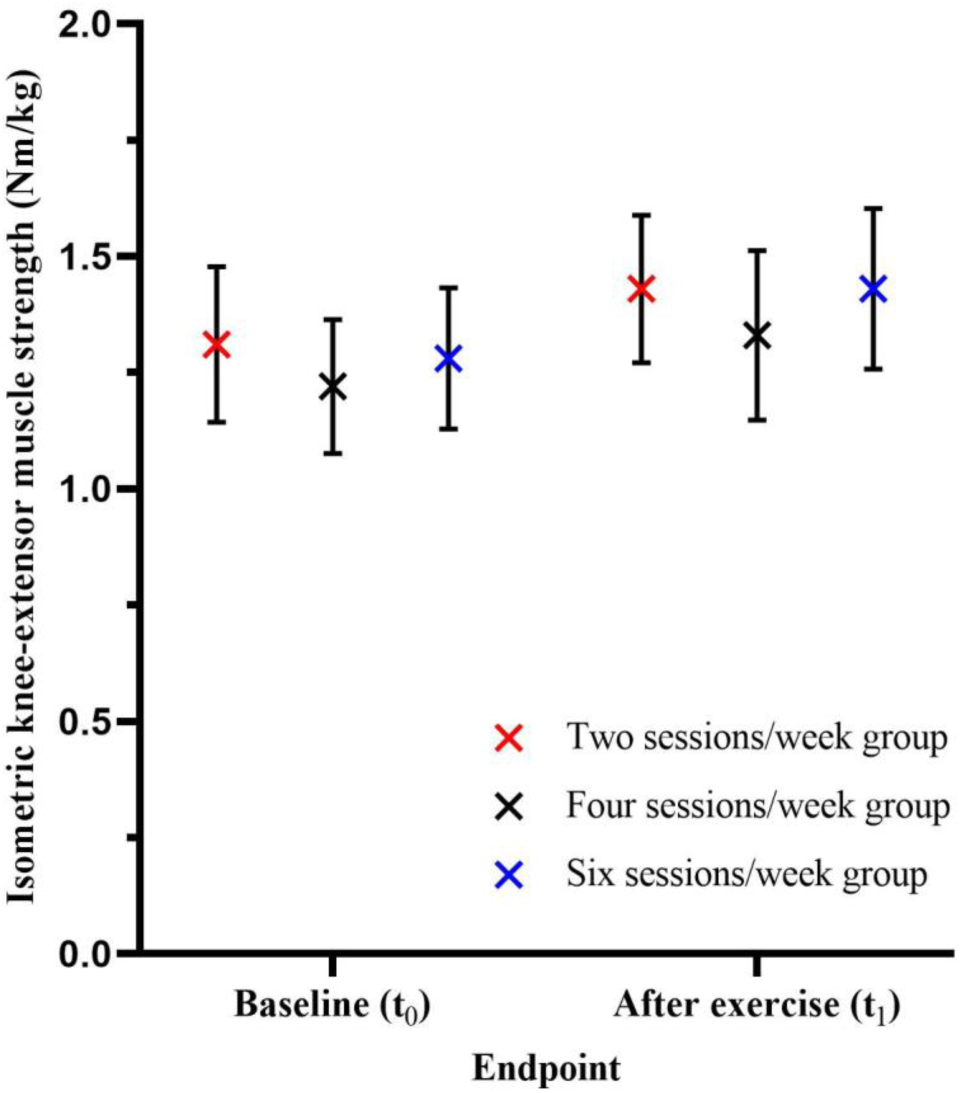
Isometric knee-extensor strength (Nm/kg) at baseline (t_0_) and after twelve weeks of home-based knee-extensor strength exercise (t_1_) across the three groups. The X represents the mean value and the whiskers the corresponding 95% confidence intervals.

**Table 2.**
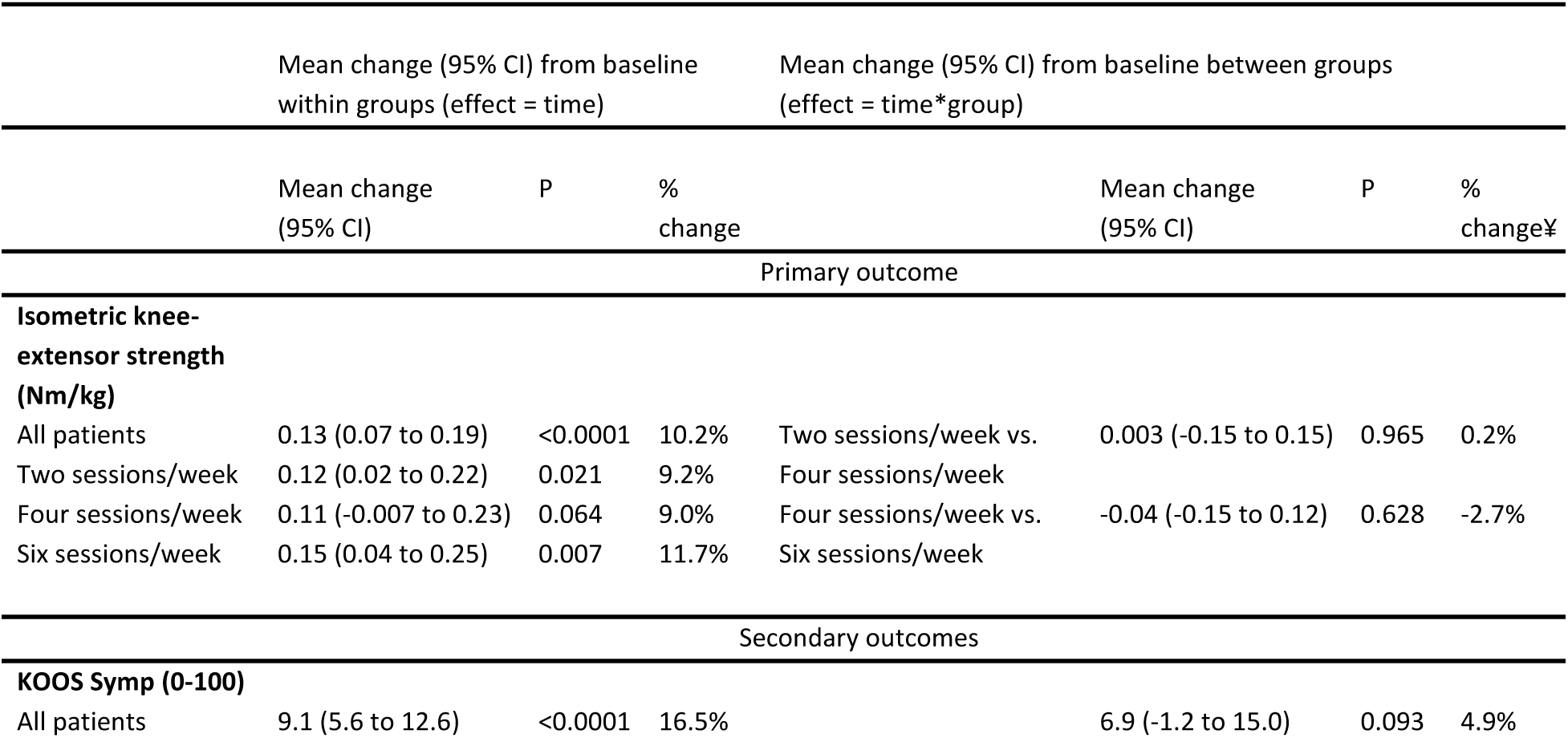
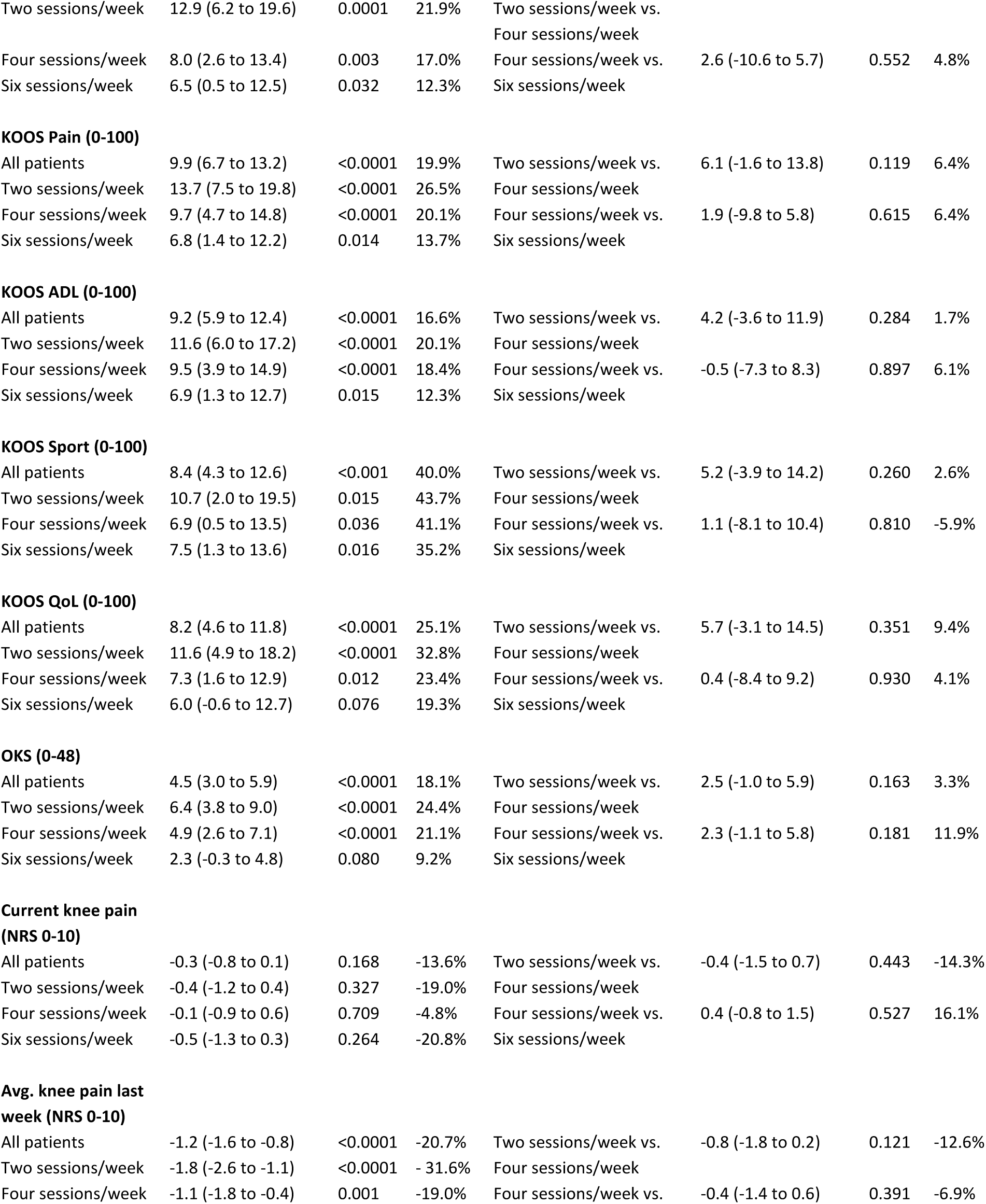
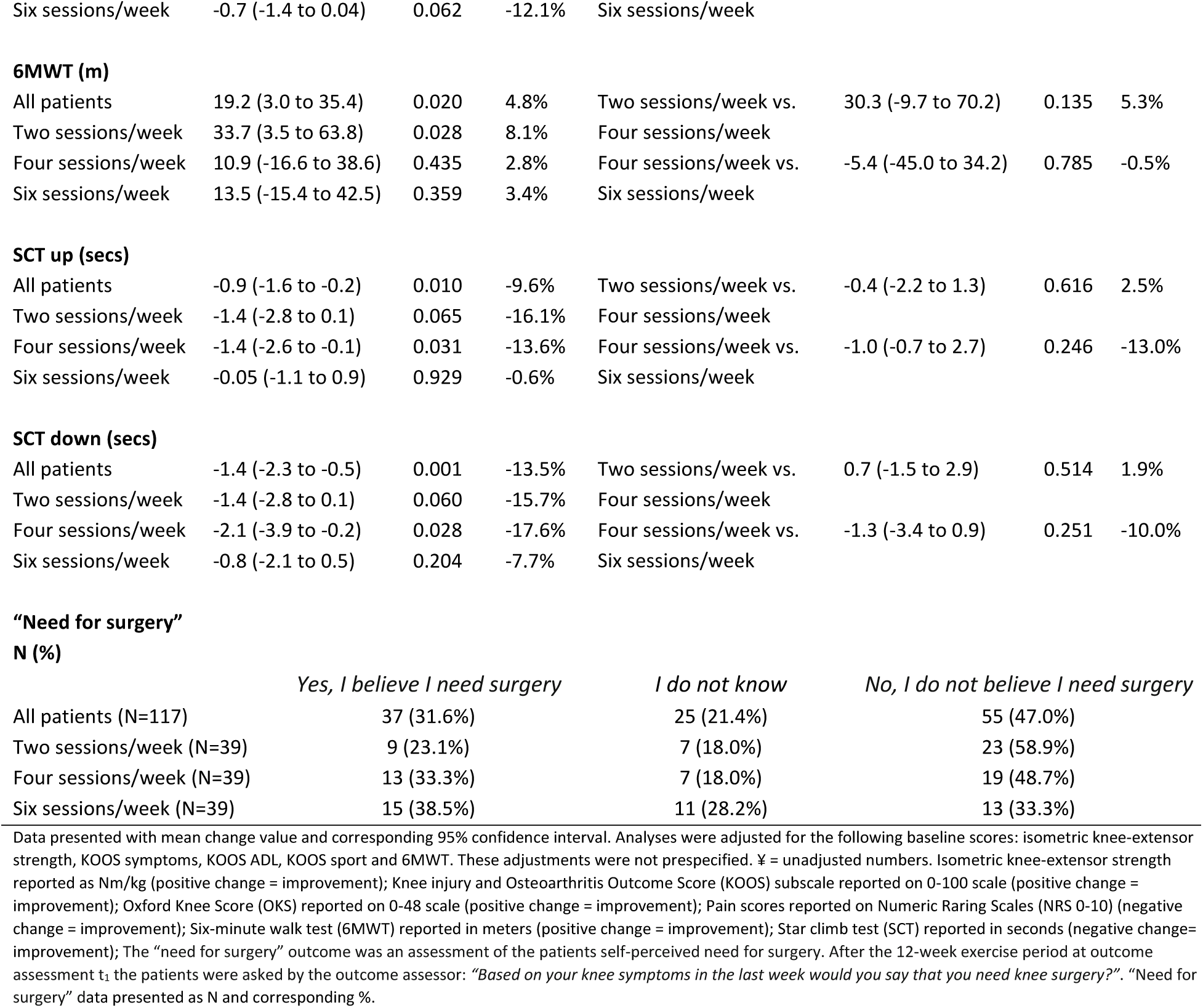
Mean change in all outcomes between baseline and following 12 weeks home-based exercise (t_0_-t_1_). Intention-to-treat analysis, N = 140. One-way ANOVA based on imputed data.

Secondary outcomes: Intention-to-treat analyses showed no between group differences for any group comparisons or secondary outcomes at the primary endpoint after 12 weeks exercise. Results from regression analyses in Supplement 4.

Secondary analysis: Intention-to-treat analyses showed statistically significant differences between the two and six sessions/week groups in favor of the two sessions/week group for *Oxford Knee Score*: 4.8 OKS points (15.2%) [1.3 to 8.3], P=0.008) and *avg. knee pain last week (NRS 0-10)*: -1.3 NRS points (−19.5%) [-2.3 to -0.2], P=0.018. No other differences were found for the secondary analysis (Supplement 3).

Due to the large proportion of patients who postponed surgery after the exercise intervention, only 32 patients were available for the post-operative intention-to-treat analyses. No between group differences for any outcomes were observed at these endpoints (Supplement 5).

### Exercise adherence

Data from 95 patients was available for the exercise adherence assessment. Of the 45 patients without available data, 23 did not complete the 12 weeks of exercise (dropped-out and missing data), 8 had less than 6 recorded exercise sessions and 14 had technical problems or lost the BandCizer© sensor. Exercise adherence was quantified as 1) *total number of sessions* and 2) *total time-under-tension (TUT)*. When exercise adherence was quantified as *total number of sessions* both the two and four sessions/week groups completed >75% of the prescribed dosage (84.8% and 81.9%, respectively). When quantified as *total time-under-tension (TUT)* no groups completed >75% of the prescribed dosage (Table 3) (Figure 3).

**Table 3.** Exercise adherence (t_0_-t_1_).

| Total number of sessions |  |  |  |  |  |  |
| --- | --- | --- | --- | --- | --- | --- |
|  | Number of prescribed sessions | Number of completed sessions (SD) | Percentage of completed sessions (SD) |  | Mean percentage difference between groups (95% CI) | P |
| All patients (N=95) | 144 | 107.6 (14.2) | 76.9% (33.6%) | Two sessions/week vs. | -2.8% | 0.741 |
| Two sessions/week (N=32) | 24 | 20.3 (7.3) | 84.8% (30.4%) | Four sessions/week | (-19.6% to 13.9%) |  |
| Four sessions/week (N=29) | 48 | 39.3 (18.4) | 81.9% (38.4%) | Four sessions/week vs. | 15.3% | 0.069 |
| Six sessions/week (N=34) | 72 | 48.0 (21.6) | 66.7% (30.1%) | Six sessions/week | (-1.3% to 31.8%) |  |
| Time-under-tension (TUT) |  |  |  |  |  |  |
|  | Prescribed TUT in secs | Completed TUT in secs (SD) | Percentage of completed TUT (SD) |  | Mean percentage difference between groups (95% CI) | P |
| All patients (N=95) | 41472 | 23412.5 (2918.5) | 56.5% (31.9%) | Two sessions/week vs. | 0.3% | 0.965 |
| Two sessions/week (N=32) | 6912 | 4477.3 (2161.9) | 64.8% (31.3%) | Four sessions/week | (-15.5% to 16.2%) |  |
| Four sessions/week (N=29) | 13824 | 9002.2 (4870.4) | 65.1% (35.2%) | Four sessions/week vs. | 17.2% | 0.031 |
| Six sessions/week (N=34) | 20736 | 9933.0 (5596.3) | 47.9% (26.9%) | Six sessions/week | (1.6% to 32.8%) |  |

Objectively quantified exercise adherence using a sensor attached to the exercise band (BandCizer© technology). Data presented with mean change value and corresponding 95% confidence interval. Statistical test: one-Way ANOVA.

**Figure 3.**
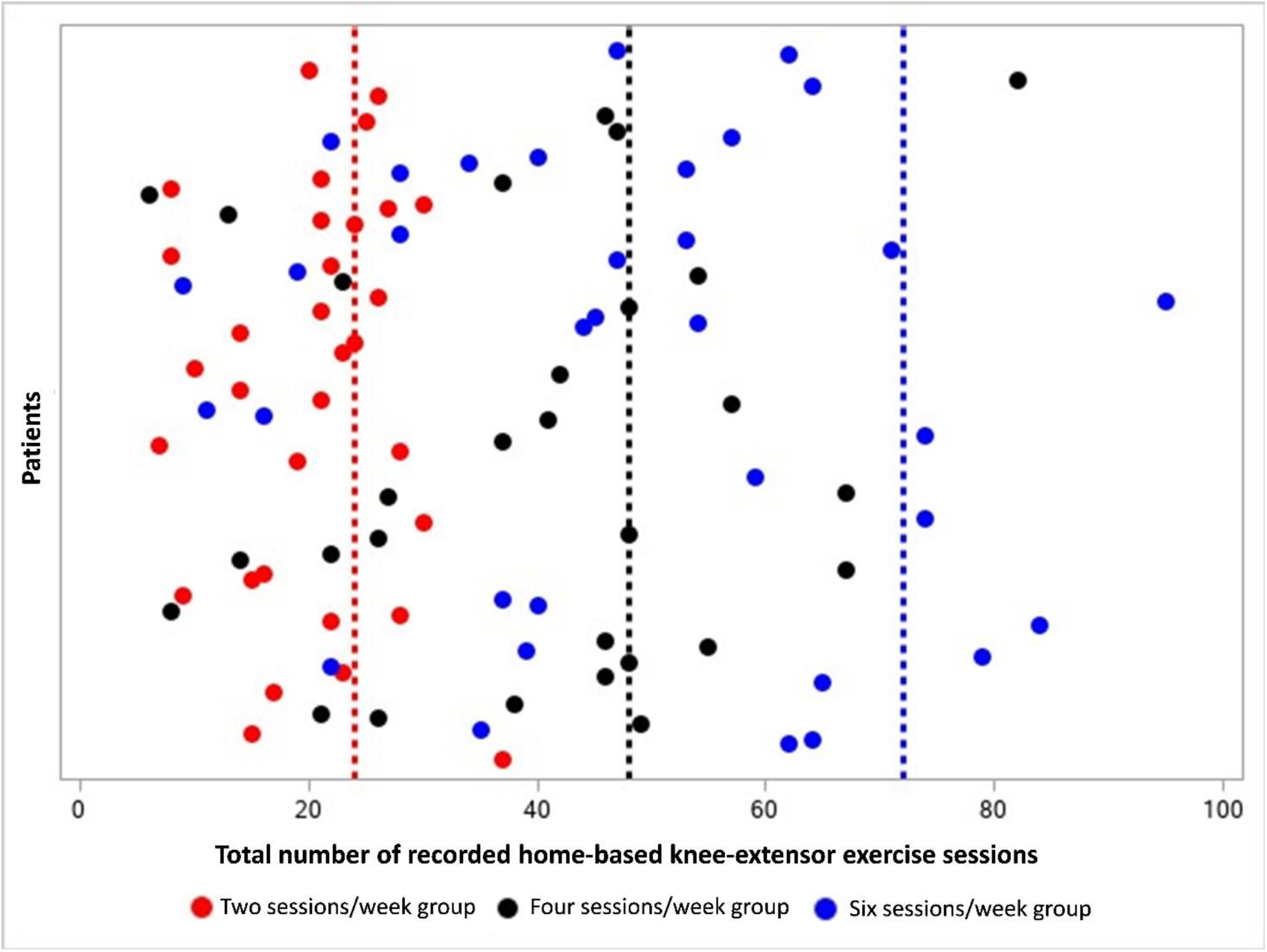
Adherence to prescribed exercise dosage across the three groups with exercise quantified as *total number of exercise sessions*. Circles represent the mean number of recorded exercise sessions for each patient. Red circles represent patients prescribed two exercise sessions per week. Black circles represent patients prescribed four exercise sessions per week. Blue circles represent patients prescribed six exercise sessions per week. The red dotted line represents the prescribed exercise dosage in the two sessions/week group (24 sessions). The black dotted line represents the prescribed exercise dosage in the four sessions/week group (48 sessions). The blue dotted line represents the prescribed exercise dosage in the six sessions/week group (72 sessions).

### Treatment decision after exercise therapy

As a post hoc analysis, the number of patients who underwent surgery and those who postponed surgery were registered. Of the 117 patients with follow-up assessments after 12 weeks of exercise (Figure 1), 79 (67.5%) postponed surgery, 32 (27.4%) underwent knee replacement, and 6 (5.1%) wanted surgery, but the orthopedic surgeon deemed this contra-indicated due to co-morbidities (Table 4) (Supplement 6).

**Table 4.**
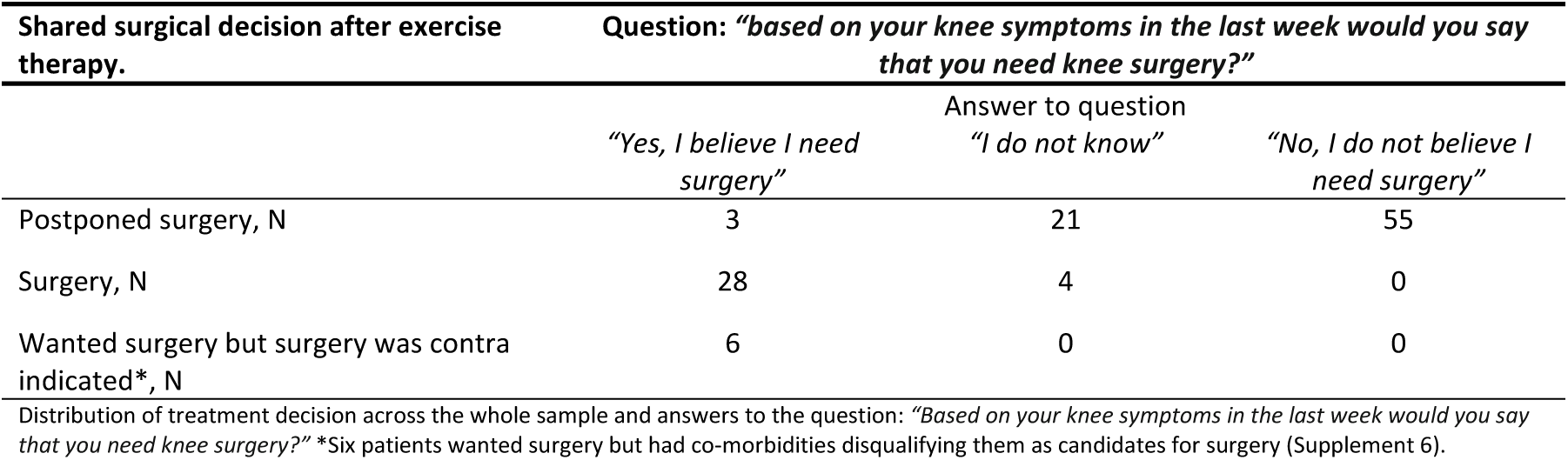
Patients’ self-perceived “need for surgery” and surgical decision after exercise therapy.

| Shared surgical decision after exercise therapy. | Question: “based on your knee symptoms in the last week would you say that you need knee surgery?” |  |  |
| --- | --- | --- | --- |
|  | Answer to question |  |  |
|  | “Yes, I believe I need surgery” | “I do not know” | “No, I do not believe I need surgery” |
| Postponed surgery, N | 3 | 21 | 55 |
| Surgery, N | 28 | 4 | 0 |
| Wanted surgery but surgery was contra indicated*, N | 6 | 0 | 0 |

Distribution of treatment decision across the whole sample and answers to the question: “Based on your knee symptoms in the last week would you say that you need knee surgery?” \*Six patients wanted surgery but had co-morbidities disqualifying them as candidates for surgery (Supplement 6).

### Harms

A total of 14 adverse events were registered during the trial period. Exacerbated knee pain due to the exercise intervention was the most frequent cause of harm (Figure 1).

## Discussion

In patients eligible for knee replacement four knee-extensor resistance exercise sessions per week were not superior to two and six sessions per week in improving isometric knee-extensor strength – indicating no exercise dose-response relationship. Independent of exercise dosage, only one in three patients completing the exercise therapy intervention decided to undergo surgery for their knee OA.

The results of the present trial are relevant for the following reasons. Firstly, larger exercise dosages do not seem to be more effective than smaller. Secondly, an exercise intervention with *one* home-based exercise can lead to clinically relevant improvements in symptoms comparable to more comprehensive interventions in patients eligible for knee replacement.^1–4,22,45,46^ Finally, a simple exercise therapy intervention, in a model of coordinated care, can prompt the majority of patients eligible for knee replacement to postpone surgery.

### Efficacy of different knee-extensor resistance exercise dosages

We found no difference in knee-extensor strength gains between the three investigated exercise dosages after twelve weeks as no between-group contrasts reached the MCID of 0.15 Nm/kg. This finding is unexpected based on the ACSM recommendations for muscle strength gains (larger exercise dosages lead to larger muscle strength gains).^33^ A possible explanation is that the ACSM recommendations are based on healthy people, not patients with knee OA eligible for knee replacement. Patients with severe knee OA likely respond differently to knee-focused exercise due to their condition and associated impairments (e.g. arthrogenic muscle inhibition)^47^ – something which could interfere with the exercise dose-response relationship classically seen in healthy people.^48^ The results suggest that patients eligible for knee replacement increase their knee-extensor strength equally when exercising with large or small dosages. This is supported by the result from our recent meta-regression analysis, in which we found no relationship between knee-extensor resistance exercise dosage and change in knee-extensor strength in patients eligible for knee replacement (meta-regression was completed after initiation of the QUADX-1 trial).^49^ Patients with knee OA might not need large exercise dosages to improve muscle strength – something also suggested in the recent START trial.^45^ In the START trial, high-intensity strength training was not superior to low-intensity strength training, nor to an attention-control in knee OA.^45^ It suggests that a classic exercise-dose-response relationship may not exist in knee OA, and that some of the effect, believed to be exercise-specific, may in fact be caused by “unspecific” or “contextual” factors.^45,50^

Another factor that may contribute to our finding of no dose-response relationship is adherence to the prescribed dosages. As seen in figure 3 there is some overlap between the completed exercise sessions across the three groups. This likely makes the difference in completed exercise between the groups less clear. Even-though the six sessions/week group completed more exercise in total, compared with the two and four sessions/week groups, the six sessions/week group had the lowest adherence relative to the prescribed dosage (66.7% of prescribed sessions), not reaching the predefined criterion of >75%. This lack of completed exercise could lead to a missing physiological response and concomitant increase in knee-extensor strength.^34,51^

Finally, the applied MCID of 0.15 Nm/kg might have been too large to establish differences between groups. However, the level of change in knee-extensor strength should also be large enough to potentially affect clinical outcomes. A meta-regression analysis from 2017 suggested that an increase of 30-40% in knee-extensor strength is needed to induce beneficial effects on pain and disability in patients with knee OA.^52^

### Secondary outcomes

For the secondary outcomes in the primary analysis, none of the differences between the groups were statistically significant or reached the MCID for any outcome. In the secondary analysis, significant differences between groups two and six sessions/week were found for OKS and avg. knee pain last week (NRS 0-10), 4.8 OKS and 1.3 NRS points, respectively, although none of the differences reached the MCID.

The two and four sessions/week groups both reached the MCID of 8-10 points for the KOOS subscales symptoms, pain and ADL, while only the two sessions/week group also reached this change for subscales sport and quality of life.^53^ The six sessions/week group did not reach the MCID for any KOOS subscales. A similar tendency was seen for OKS and 6MWT where the two sessions/week group reached the MCID of 6 points and 20 m, respectively, while the four and six sessions/week groups did not.^54,55^ On the NRS 0-10 scale for pain the two and four sessions/week groups both reached ‘slightly better’ improvements, with - 1.8 and -1.1 changes in avg. knee pain last week (NRS 0-10), respectively, while the six sessions/week did not.^56^ Finally, for the SCT no MCID is known while the minimal detectable change is reported to be 2.6 seconds.^57^ No groups reached this for neither the up or down stair climbing assessment.

In general, the two and four sessions/week groups reached the MCID for the outcomes more often than the six sessions/week group. This could be explained by the larger exercise dose with more frequent sessions leaving less time to recover between sessions – something that could lead to increase in knee pain and decreased physical function.

### Implications for *one* home-based exercise and coordinated non-surgical and surgical care

The results from the QUADX-1 trial are comparable to other trials reporting similar proportions of patients with severe OA who postpone surgery and corresponding clinically relevant improvements in patient-reported outcomes after exercise therapy.^1–4^ Compared to the intervention in the QUADX-1 trial, the exercise therapy interventions in these trials are more comprehensive and costly, comprising more exercises and supervision. This suggests that the intervention and associated exercise dosage needed to improve symptoms in patients eligible for knee replacement does not have to be extensive or comprehensive.^45^ This corresponds well to the results from the supplementary regression analyses and our recent meta-regression analysis indicating no dose-response between exercise dosage and change in outcomes before scheduled knee replacement.^49^ A minimal exercise approach as part of coordinated non-surgical and surgical care pathway in severe knee OA seems relevant based on current dose-response findings^49^ and specific exercise effects in knee OA.^50^ The effects observed in the present trial across different outcomes may be a small specific effect of exercise and/or of other contextual factors. It could be caused by contact with healthcare professionals^58–60^, regression to the mean, natural cause of the disease,^61^ or simply by placebo effect^50^ – making it difficult to ascribe too much specific cause-of-effect to exercise therapy. In line with this, the recent DISCO trial found equivalent improvements after a supervised exercise and education program, and saline injections in patients with knee OA.^50^ These factors question the best way to provide (exercise) care for patients with knee OA and could suggest that supervised exercise is not the most cost-effective approach.

The large number of patients who postponed surgery highlights the importance of coordinating non-surgical and surgical care in patients eligible for knee replacement. The proportion of patients postponing and choosing surgery across the three groups appeared similar – indicating that the decision to postpone surgery was independent of the prescribed exercise therapy dosage (not powered for this outcome). A contributing factor explaining the large number of patients postponing surgery could be the non-specific effect of the applied ‘model’ of coordinated non-surgical and surgical care.^16,59,60,62–64^ In this ‘model’, the patients’ decision on surgical treatment was re-evaluated by the patient and orthopedic surgeon after the exercise period. This re-evaluation based on symptom changes, combined with additional attention from an orthopedic surgeon, could have facilitated the patients’ decision to postpone surgery.^58–60,65–68^ This is exemplified in table 4 showing that patients who believe they need surgery, undergo surgery, while those who “don’t know” or do not believe they need surgery postpone it.

In the Enhanced Recovery After Surgery (ERAS) concept it is assumed that exercise therapy before planned surgery (exercise-based pre-habilitation) is always followed by surgery.^69–73^ We have previously argued that the premise for exercise therapy before potential surgery – to enhance post-surgical outcomes in patients eligible for knee replacement – should be questioned^49^ as several systematic reviews conclude no clinically-relevant effect post-operatively.^49,74–83^ Instead of being a predetermined care pathway (leading to surgery), exercise therapy before potential surgery could be used to inform the shared decision-making process when planning a care pathway,^8,17,18^ which complies with guideline recommendations while being cost-effective.^5–10,84^ Based on the results from the QUADX-1 trial, we suggest using simple (*one* exercise) home-based resistance exercise therapy within the ERAS concept to “pre-evaluate” the need for surgery in patients with severe knee OA rather than to “prepare” patients for surgery.

### Limitations

The Danish healthcare system is publicly funded, and treatment is therefore free. Refusing surgery after having been on a waiting list does not postpone the possibility of surgery for years. The patients can be re-assessed by an orthopaedic surgeon within months and have surgery scheduled if needed. This might limit the comparability to other countries with a different healthcare system. The patients were aware that the sensor attached to the exercise band recorded their exercise adherence. This might potentially have affected their exercise adherence.^85^

## Conclusion

In patients eligible for knee-replacement we found no between-group differences in isometric knee extensor strength after 2, 4 and 6 knee-extensor resistance exercise sessions per week. We saw no indication of an exercise dose-response relationship for isometric knee-extensor strength and only clinically irrelevant within group changes. For some secondary outcome (e.g. KOOS subscales) we found clinically relevant within group changes, which could help explain why only one in three patients decided to have surgery after the simple home-based exercise intervention.

## Data Availability

The raw data is available by contacting the corresponding author Rasmus Skov Husted via

## Acknowledgements

Acknowledgement go to: Research assistant Line Holst for help with trial management and outcomes assessment, and to the physiotherapists in the participating municipalities (Broendby, Hvidovre and Copenhagen) and the orthopedic surgeons at the Orthopedic Department at Copenhagen University Hospital Amager-Hvidovre.

## Contributions

Conception and design: Husted, Troelsen, Rathleff, Thorborg and Bandholm.

Data acquisition: Husted and Grønfeldt.

Analysis and interpretation of the data: All authors.

Drafting of the article: Husted.

Critical revision of the article for important intellectual content: All authors.

Statistical expertise: Kallemose.

Final approval of the article: All authors. Mr. Husted and Prof. Bandholm takes responsibility for the integrity of the work as a whole.

## Supplement 1. Missing data at the different endpoints

**eTable 1.** Missing data at the different endpoints.

| Outcomes | Baseline (t<sub>0</sub>) |  |  |  |
| --- | --- | --- | --- | --- |
|  | All patients<br>N = 140 | Two sessions/week<br>N = 47 | Four sessions/week<br>N = 47 | Six sessions/week<br>N = 46 |
| Gender (f/m) | - | - | - | - |
| Age (years) | - | - | - | - |
| Weight (kg) | - | - | - | - |
| Height (cm) | - | - | - | - |
| Municipality (Cph/Hvi/Bro) | - | - | - | - |
| Kellgren and Lawrence score (2/3/4) | 2 | 1 | 1 | - |
| Current knee pain (NRS 0-10) | - | - | - | - |
| Avg. knee pain last week (NRS 0-10) | - | - | - | - |
| Isometric knee-extensor strength (Nm/kg) | - | - | - | - |
| KOOS Symp (0-100) | - | - | - | - |
| KOOS Pain (0-100) | - | - | - | - |
| KOOS ADL (0-100) | - | - | - | - |
| KOOS Sport (0-100) | - | - | - | - |
| KOOS QoL (0-100) | 1 | - | 1 | - |
| OKS (0-48) | - | - | - | - |
| 6MWT (m) | 3 | 1 | 1 | 1 |
| SCT up (secs) | 2 | - | 1 | 1 |
| SCT down (secs) | 2 | - | 1 | 1 |

| Outcomes | Following 12 weeks home-based exercise (t<sub>1</sub>) |  |  |  |
| --- | --- | --- | --- | --- |
|  | All patients<br>N = 140 | Two sessions/week<br>N = 47 | Four sessions/week<br>N = 47 | Six sessions/week<br>N = 46 |
| Current knee pain (NRS 0-10) | 23 | 8 | 8 | 7 |
| Avg. knee pain last week (NRS 0-10) | 23 | 8 | 8 | 7 |
| Isometric knee-extensor strength (Nm/kg) | 23 | 8 | 8 | 7 |
| KOOS Symp (0-100) | 23 | 8 | 8 | 7 |
| KOOS Pain (0-100) | 23 | 8 | 8 | 7 |
| KOOS ADL (0-100) | 23 | 8 | 8 | 7 |
| KOOS Sport (0-100) | 23 | 8 | 8 | 7 |
| KOOS QoL (0-100) | 23 | 8 | 8 | 7 |
| OKS (0-48) | 23 | 8 | 8 | 7 |
| 6MWT (m) | 29 | 11 | 9 | 9 |
| SCT up (secs) | 31 | 11 | 11 | 9 |
| SCT down (secs) | 31 | 11 | 11 | 9 |

| Outcomes | Acutely after surgery (t<sub>2</sub>) |  |  |  |
| --- | --- | --- | --- | --- |
|  | All patients<br>N = 32 | Two sessions/week<br>N = 7 | Four sessions/week<br>N = 12 | Six sessions/week<br>N = 13 |
| Current knee pain (NRS 0-10) | 15 | 4 | 9 | 2 |
| Isometric knee-extensor strength (Nm/kg) | 16 | 4 | 9 | 3 |
| 6MWT (m) | 26 | 5 | 12 | 9 |
| SCT up (secs) | 28 | 5 | 12 | 11 |
| SCT down (secs) | 28 | 5 | 12 | 11 |

| Three months after surgery (t<sub>3</sub>) |  |  |  |  |
| --- | --- | --- | --- | --- |
| Outcomes | All patients<br>N = 32 | Two sessions/week<br>N = 7 | Four sessions/week<br>N = 12 | Six sessions/week<br>N = 13 |
| Current knee pain (NRS 0-10) | 7 | 2 | 4 | 1 |
| Avg. knee pain last week (NRS 0-10) | 7 | 2 | 4 | 1 |
| Isometric knee-extensor strength (Nm/kg) | 8 | 2 | 4 | 2 |
| KOOS Symp (0-100) | 7 | 2 | 4 | 1 |
| KOOS Pain (0-100) | 7 | 2 | 4 | 1 |
| KOOS ADL (0-100) | 7 | 2 | 4 | 1 |
| KOOS Sport (0-100) | 7 | 2 | 4 | 1 |
| KOOS QoL (0-100) | 7 | 2 | 4 | 1 |
| OKS (0-48) | 7 | 2 | 4 | 1 |
| 6MWT (m) | 9 | 2 | 5 | 2 |
| SCT up (secs) | 10 | 2 | 6 | 2 |
| SCT down (secs) | 10 | 2 | 6 | 2 |

Appendix 1. Cph = Copenhagen, Hvi = Hvidovre, Bro = Broendby, KOOS = Knee injury and Osteoarthritis Outcome Score, OKS = Oxford Knee Score, 6MWT = six-minute walk test, SCT = Stair climb test. Group A = 2 exercise sessions/week, Group B = 4 exercise sessions/week, Group C = 6 exercise sessions/week.

## Supplement 2. Separate baseline characteristics for the patients lost to follow-up, the remaining patients and the sub-sample that underwent surgery

**eTable 2.** Baseline characteristics (t_0_). Participants lost to follow-up.

| Characteristics | All patients | Two sessions/week | Four sessions/week | Six sessions/week |
| --- | --- | --- | --- | --- |
| Mean (SD) | N = 23 | N = 8 | N = 8 | N = 7 |
| Gender (f/m) | 14/9 | 4/4 | 7/1 | 3/4 |
| Age (years) | 65.7 (9.6) | 68.8 (8.9) | 69.6 (9.9) | 57.8 (5.1) |
| Weight (kg) | 93.9 (16.9) | 100.2 (17.0) | 88.3 (14.9) | 93.1 (18.9) |
| Height (cm) | 167.9 (7.4) | 166.3 (8.0) | 166.3 (5.3) | 171.9 (8.3) |
| Municipality (Cph/Hvi/Bro) | 16/6/1 | 5/3/0 | 6/1/1 | 5/2/0 |
| Kellgren and Lawrence score (2/3/4) | 5/13/5 | 2/4/2 | 2/3/3 | 1/6/0 |
| Current knee pain (NRS 0-10) | 2.1 (2.5) | 2.3 (3.1) | 1.8 (2.4) | 2.3 (2.1) |
| Avg. knee pain last week (NRS 0-10) | 5.9 (1.7) | 6.3 (1.6) | 5.6 (1.7) | 5.7 (1.9) |
| Isometric knee-extensor strength (Nm/kg) | 1.28 (0.55) | 1.19 (0.61) | 1.21 (0.36) | 1.46 (0.69) |
| KOOS Symp (0-100) | 51.6 (18.5) | 52.7 (23.4) | 50.9 (13.6) | 51.0 (19.9) |
| KOOS Pain (0-100) | 46.4 (15.3) | 46.3 (22.2) | 49.7 (8.6) | 42.9 (12.5) |
| KOOS ADL (0-100) | 50.6 (13.8) | 51.7 (18.7) | 49.8 (8.6) | 50.2 (14.0) |
| KOOS Sport (0-100) | 20.0 (19.8) | 28.8 (27.5) | 15.0 (11.9) | 15.0 (13.5) |
| KOOS QoL (0-100) | 30.7 (16.9) | 36.7 (19.9) | 28.1 (14.9) | 26.8 (16.0) |
| OKS (0-48) | 22.8 (7.4) | 23.3 (9.1) | 22.5 (7.1) | 22.7 (6.8) |
| 6MWT (m) | 403.4 (92.8) | 383.4 (93.6) | 388.7 (80.3) | 438.2 (109.2) |
| SCT up (secs) | 8.4 (2.8) | 8.9 (2.8) | 9.6 (2.9) | 6.5 (1.7) |
| SCT down (secs) | 8.9 (3.5) | 9.1 (2.3) | 10.7 (4.4) | 6.7 (2.9) |

Cph = Copenhagen, Hvi = Hvidovre, Bro = Broendby, NRS = Numeric Rating Scale, KOOS = Knee injury and Osteoarthritis Outcome Score, OKS = Oxford Knee Score, 6MWT = six-minute walk test, SCT = Stair climb test.

**eTable 3.** Baseline characteristics (t_0_). Remaining participants.

| Characteristics | All patients | Two sessions/week | Four sessions/week | Six sessions/week |
| --- | --- | --- | --- | --- |
| Mean (SD) | N = 117 | N = 39 | N = 39 | N = 39 |
| Gender (f/m) | 62/55 | 18/21 | 22/17 | 22/17 |
| Age (years) | 66.9 (9.9) | 67.2 (10.1) | 66.4 (9.9) | 67.2 (10.1) |
| Weight (kg) | 91.5 (20.5) | 90.5 (16.9) | 94.7 (23.1) | 89.2 (20.9) |
| Height (cm) | 169.5 (8.5) | 169.2 (6.8) | 170.7 (8.1) | 168.6 (10.3) |
| Municipality (Cph/Hvi/Bro) | 58/38/21 | 17/13/9 | 17/15/7 | 24/10/5 |
| Kellgren and Lawrence score (2/3/4) | 15/48/52 | 3/16/19 | 7/16/15 | 5/16/18 |
| Current knee pain (NRS 0-10) | 2.2 (2.2) | 2.1 (2.3) | 2.2 (2.1) | 2.4 (2.2) |
| Avg. knee pain last week (NRS 0-10) | 5.8 (1.5) | 5.6 (1.6) | 5.9 (1.7) | 5.8 (1.4) |
| Isometric knee-extensor strength (Nm/kg) | 1.26 (0.53) | 1.33 (0.57) | 1.21 (0.55) | 1.24 (0.48) |
| KOOS Symp (0-100) | 55.7 (18.9) | 60.2 (18.8) | 53.6 (17.4) | 53.2 (20.1) |
| KOOS Pain (0-100) | 50.4 (16.8) | 52.8 (15.4) | 47.5 (18.1) | 50.8 (16.1) |
| KOOS ADL (0-100) | 56.2 (17.9) | 58.9 (16.8) | 52.2 (18.9) | 57.4 (17.9) |
| KOOS Sport (0-100) | 21.3 (20.9) | 23.6 (23.3) | 17.7 (18.2) | 22.4 (21.2) |
| KOOS QoL (0-100) | 33.1 (16.3) | 35.1 (15.9) | 32.2 (16.8) | 31.9 (16.2) |
| OKS (0-48) | 25.2 (7.6) | 26.9 (7.0) | 23.3 (8.4) | 25.3 (7.2) |
| 6MWT (m) | 425.1 (96.5) | 440.5 (86.3) | 412.3 (109.1) | 421.2 (93.9) |
| SCT up (secs) | 9.0 (4.4) | 7.8 (3.0) | 9.9 (5.3) | 9.1 (4.3) |
| SCT down (secs) | 10.0 (6.2) | 8.3 (4.8) | 11.1 (6.9) | 10.7 (6.5) |

Cph = Copenhagen, Hvi = Hvidovre, Bro = Broendby, NRS = Numeric Rating Scale, KOOS = Knee injury and Osteoarthritis Outcome Score, OKS = Oxford Knee Score, 6MWT = six-minute walk test, SCT = Stair climb test.

**eTable 4.** Baseline characteristics for the sub-sample that underwent surgery (t_0_).

| Characteristics | All patients | Two sessions/week | Four sessions/week | Six sessions/week |
| --- | --- | --- | --- | --- |
| Mean (SD) | N = 32 | N = 7 | N = 12 | N = 13 |
| Gender (f/m) | 20/12 | 6/1 | 8/4 | 6/7 |
| Age (years) | 66.4 (9.0) | 68.8 (12.7) | 65.2 (8.3) | 66.2 (7.9) |
| Weight (kg) | 93.2 (16.6) | 90.5 (16.4) | 92.2 (17.0) | 95.7 (17.2) |
| Height (cm) | 170.6 (9.6) | 165.7 (9.4) | 169.9 (7.5) | 173.9 (10.7) |
| Municipality (Cph/Hvi/Bro) | 16/10/6 | 4/2/1 | 3/6/3 | 9/2/2 |
| Kellgren and Lawrence score (2/3/4) | 4/10/18 | 2/2/3 | 1/4/7 | 1/4/8 |
| Current knee pain (NRS 0-10) | 2.3 (2.0) | 1.9 (1.3) | 3.0 (2.1) | 2.0 (2.2) |
| Avg. knee pain last week (NRS 0-10) | 6.1 (1.3) | 5.4 (1.0) | 6.9 (1.0) | 5.8 (1.4) |
| Isometric knee-extensor strength (Nm/kg) | 1.18 (0.51) | 1.02 (0.43) | 1.10 (0.60) | 1.35 (0.43) |
| KOOS Symp (0-100) | 52.3 (17.8) | 61.2 (16.3) | 45.8 (15.7) | 53.6 (19.2) |
| KOOS Pain (0-100) | 43.6 (14.5) | 47.6 (15.7) | 37.0 (16.0) | 47.4 (10.8) |
| KOOS ADL (0-100) | 51.0 (14.8) | 54.8 (14.1) | 42.6 (15.8) | 56.7 (11.3) |
| KOOS Sport (0-100) | 16.9 (17.4) | 12.9 (16.3) | 13.3 (18.0) | 22.3 (17.4) |
| KOOS QoL (0-100) | 27.1 (15.4) | 28.6 (16.5) | 22.4 (15.6) | 30.8 (14.8) |
| OKS (0-48) | 22.6 (7.3) | 24.9 (7.0) | 18.5 (7.1) | 25.1 (6.4) |
| 6MWT (m) | 408.0 (96.9) | 428.6 (90.8) | 372.7 (108.4) | 429.5 (85.8) |
| SCT up (secs) | 9.4 (4.8) | 7.9 (2.6) | 11.2 (5.6) | 8.6 (4.6) |
| SCT down (secs) | 10.8 (8.2) | 8.3 (3.1) | 13.4 (10.4) | 9.7 (7.6) |
| TKA/UKA | 20/12 | 5/2 | 7/5 | 8/5 |

Cph = Copenhagen, Hvi = Hvidovre, Bro = Broendby, KOOS = Knee injury and Osteoarthritis Outcome Score, OKS = Oxford Knee Score, 6MWT = six-minute walk test, SCT = Stair climb test, TKA = Total Knee Arthroplasty, UKA = Unicompartmental Knee Arthroplasty.

## Supplement 3. Secondary analyses (Mean change between group two and six sessions/week)

**eTable 5.** Mean change between group two and six sessions/week for all outcomes between baseline and following 12 weeks home-based exercise (t_0_-t_1_). Intention-to-treat analysis, N = 140. One-way ANOVA based on imputed data.

| Mean change (95% CI) from baseline between groups (effect = time*group)<br>Two sessions/week vs. Six sessions/week | Mean change (95% CI) | P | % change¥ |
| --- | --- | --- | --- |
| Primary outcome |  |  |  |
| Isometric knee-extensor strength (Nm/kg) | -0.03 (-0.19 to 0.12) | 0.654 | -2.6% |
| Secondary outcomes |  |  |  |
| KOOS Symp (0-100) | 9.4 (1.1 to 17.7) | 0.027 | 9.6% |
| KOOS Pain (0-100) | 8.1 (0.1 to 16.1) | 0.046 | 12.8% |
| KOOS ADL (0-100) | 4.7 (-3.1 to 12.5) | 0.234 | 7.8% |
| KOOS Sport (0-100) | 4.1 (-5.1 to 13.2) | 0.376 | 8.5% |
| KOOS QoL (0-100) | 5.3 (-3.5 to 14.1) | 0.232 | 13.5% |
| OKS (0-48) | 4.8 (1.3 to 8.3) | 0.008 | 15.2% |
| Current knee pain (NRS 0-10) | 0.1 (-1.1 to 1.2) | 0.901 | 1.8% |
| Avg. knee pain last week (NRS 0-10) | -1.3 (-2.3 to -0.2) | 0.018 | -19.5% |
| 6MWT (m) | 24.9 (-14.9 to 64.7) | 0.218 | 4.7% |
| SCT up (secs) | -1.5 (-3.2 to 0.3) | 0.095 | -15.5% |
| SCT down (secs) | -0.8 (-2.8 to 1.3) | 0.468 | -8.0% |

Data presented with mean change value and corresponding 95% confidence interval. Analyses were adjusted for the following baseline scores: isometric knee-extensor strength, KOOS symptoms, KOOS ADL, KOOS sport and 6MWT. These adjustments were not prespecified. ¥ = unadjusted numbers. Isometric knee-extensor strength reported as Nm/kg (positive change = improvement); Knee injury and Osteoarthritis Outcome Score (KOOS) subscale reported on 0-100 scale (positive change = improvement); Oxford Knee Score (OKS) reported on 0-48 scale (positive change = improvement); Pain scores reported on Numeric Rating Scales (NRS 0-10) (negative change = improvement); Six-minute walk test (6MWT) reported in meters (positive change = improvement); Star climb test (SCT) reported in seconds (negative change = improvement).

## Supplement 4. Supplementary regression analyses

For the supplementary regression analyses the three groups were pooled into one sample. No association was observed between the level of exercise adherence and pre-operative changes for any outcomes, except for a weak inverse association between *total number of sessions* and change in the *six-minute walk test* (Slope -0.7323 [95% -1.819 to -0.1826]) (eTable 6).

**eTable 6.** Simple regression models.

| Independent variable | Dependent variable - Change scores from baseline to after 12 weeks of exercise (t<sub>0</sub>-t<sub>1</sub>) | Slope | 95% CI |
| --- | --- | --- | --- |
| **Exercise adherence quantified as *total number of sessions*** |  |  |  |
|  | Knee-extensor muscle strength (Nm/kg) | -0.0015 | -0.0037 to 0.0008 |
|  | KOOS Symptoms | -0.0160 | -0.1877 to 0.1557 |
|  | KOOS Pain | 0.0078 | -0.1532 to 0.1687 |
|  | KOOS ADL | 0.0556 | -0.1105 to 0.2218 |
|  | KOOS Sport | 0.0056 | -0.2013 to 0.2125 |
|  | KOOS QoL | -0.0375 | -0.2280 to 0.1529 |
|  | OKS | -0.0256 | -0.1065 to 0.0552 |
|  | Current pain (0-10 NRS) | -0.0165 | -0.0405 to 0.0075 |
|  | Avg. pain last week (0-10 NRS) | -0.0042 | -0.0282 to 0.0197 |
|  | SMWT (meters) | -0.7323 | -1.2819 to -0.1826 |
|  | SCT up (seconds) | 0.0174 | -0.0097 to 0.0444 |
|  | SCT down (seconds) | 0.0026 | -0.0367 to 0.0419 |
| **Exercise adherence quantified as *total time-under-tension (TUT)*** |  |  |  |
|  | Knee-extensor muscle strength (Nm/kg) | -0.000006 | -0.00001 to 0.000003 |
|  | KOOS Symptoms | 0.000007 | -0.0007 to 0.0008 |
|  | KOOS Pain | -0.00009 | -0.0007 to 0.0006 |
|  | KOOS ADL | 0.00004 | -0.0006 to 0.0007 |
|  | KOOS Sport | -0.00004 | -0.0009 to 0.0008 |
|  | KOOS QoL | -0.0004 | -0.0011 to 0.0004 |
|  | OKS | -0.0002 | -0.0005 to 0.0002 |
|  | Current pain (0-10 NRS) | -0.00004 | -0.0001 to 0.00005 |
|  | Avg. pain last week (0-10 NRS) | 0.00002 | -0.00008 to 0.0001 |
|  | SMWT (meters) | -0.0022 | -0.0044 to 0.00006 |
|  | SCT up (seconds) | 0.00001 | -0.00009 to 0.0001 |
|  | SCT down (seconds) | -0.00007 | -0.0002 to 0.00008 |

TUT = Time-under-tension, KOOS = Knee injury and Osteoarthritis Outcome Score, OKS = Oxford Knee Score, 6MWT = Six-minute walk test, SCT = Stair climb test.

## Supplement 5. Results for the post-surgical assessments (t_2_ and t_3_)

**Primary analyses (t**_**0**_**-t**_**2**_)

**eTable 7.**
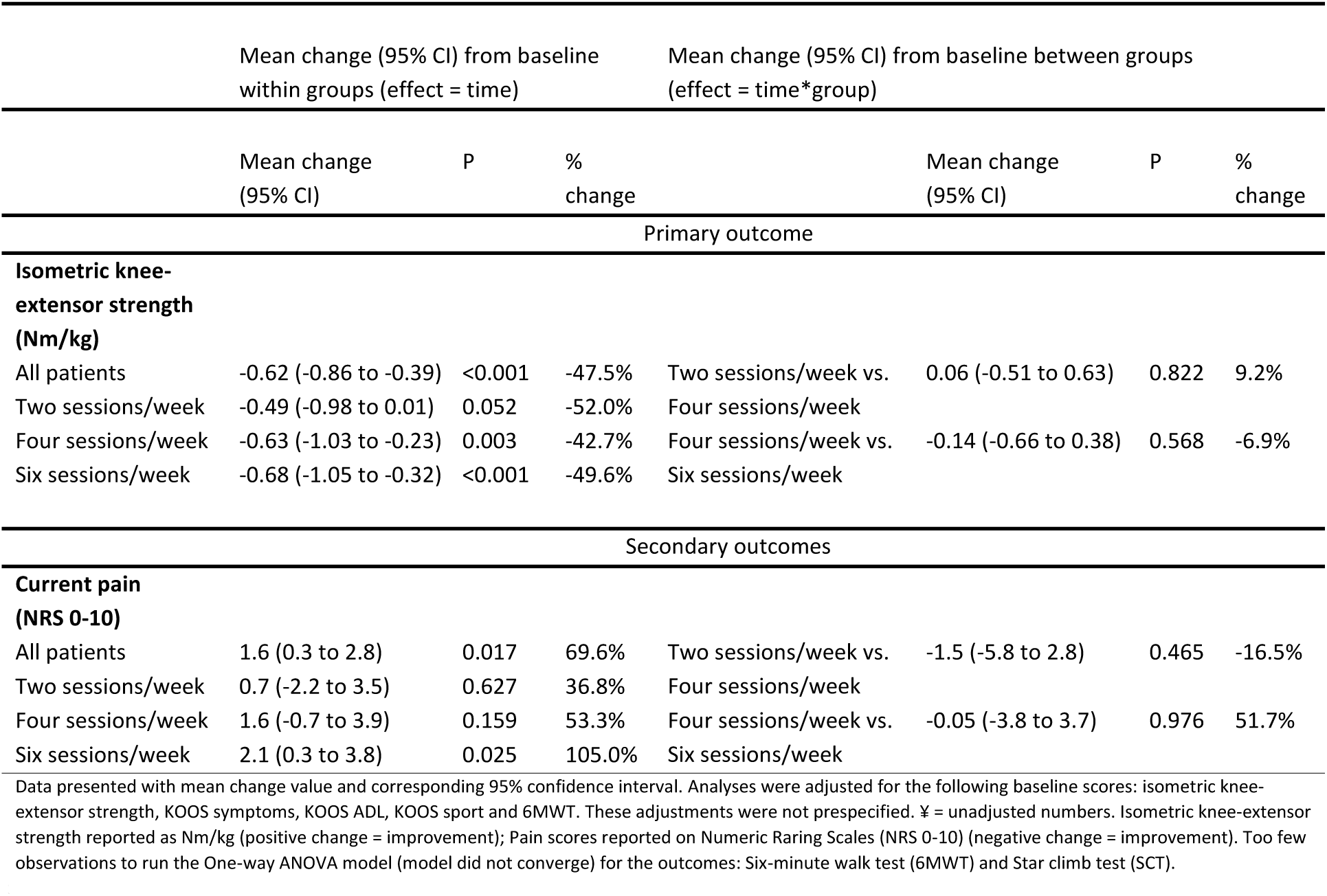
Mean change in all outcomes between baseline and acutely after surgery (t_0_-t_2_). Intention-to-treat analysis, N = 32. One-way ANOVA based on imputed data.

**Secondary analyses (Mean change between group two and six sessions/week)**

**eTable 8.**
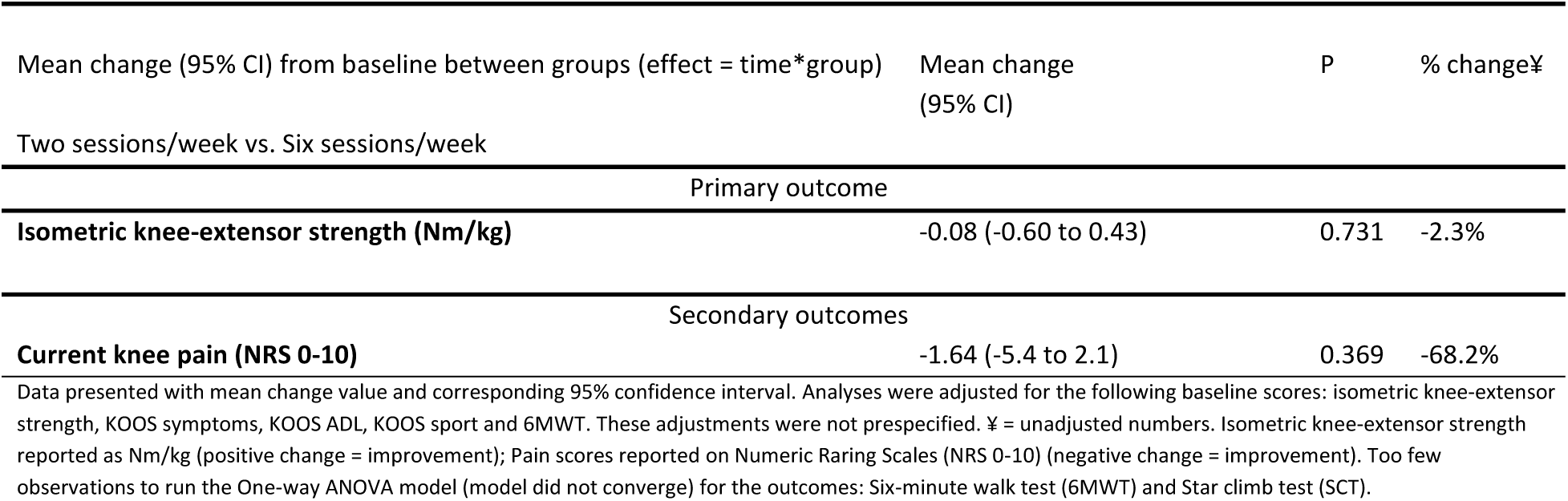
Mean change between group two and six sessions/week for all outcomes between baseline and acutely after surgery (t_0_-t_2_). Intention-to-treat analysis, N = 32. One-way ANOVA based on imputed data.

| Mean change (95% CI) from baseline between groups (effect = time*group) | Mean change (95% CI) | P | % change¥ |
| --- | --- | --- | --- |
| Two sessions/week vs. Six sessions/week |  |  |  |
| Primary outcome |  |  |  |
| **Isometric knee-extensor strength (Nm/kg)** | -0.08 (-0.60 to 0.43) | 0.731 | -2.3% |
| Secondary outcomes |  |  |  |
| **Current knee pain (NRS 0-10)** | -1.64 (-5.4 to 2.1) | 0.369 | -68.2% |

Data presented with mean change value and corresponding 95% confidence interval. Analyses were adjusted for the following baseline scores: isometric knee-extensor strength, KOOS symptoms, KOOS ADL, KOOS sport and 6MWT. These adjustments were not prespecified. ¥ = unadjusted numbers. Isometric knee-extensor strength reported as Nm/kg (positive change = improvement); Pain scores reported on Numeric Rating Scales (NRS 0-10) (negative change = improvement). Too few observations to run the One-way ANOVA model (model did not converge) for the outcomes: Six-minute walk test (6MWT) and Star climb test (SCT).

**Primary analyses (t**_**0**_**-t**_**3**_)

**eTable 9.**
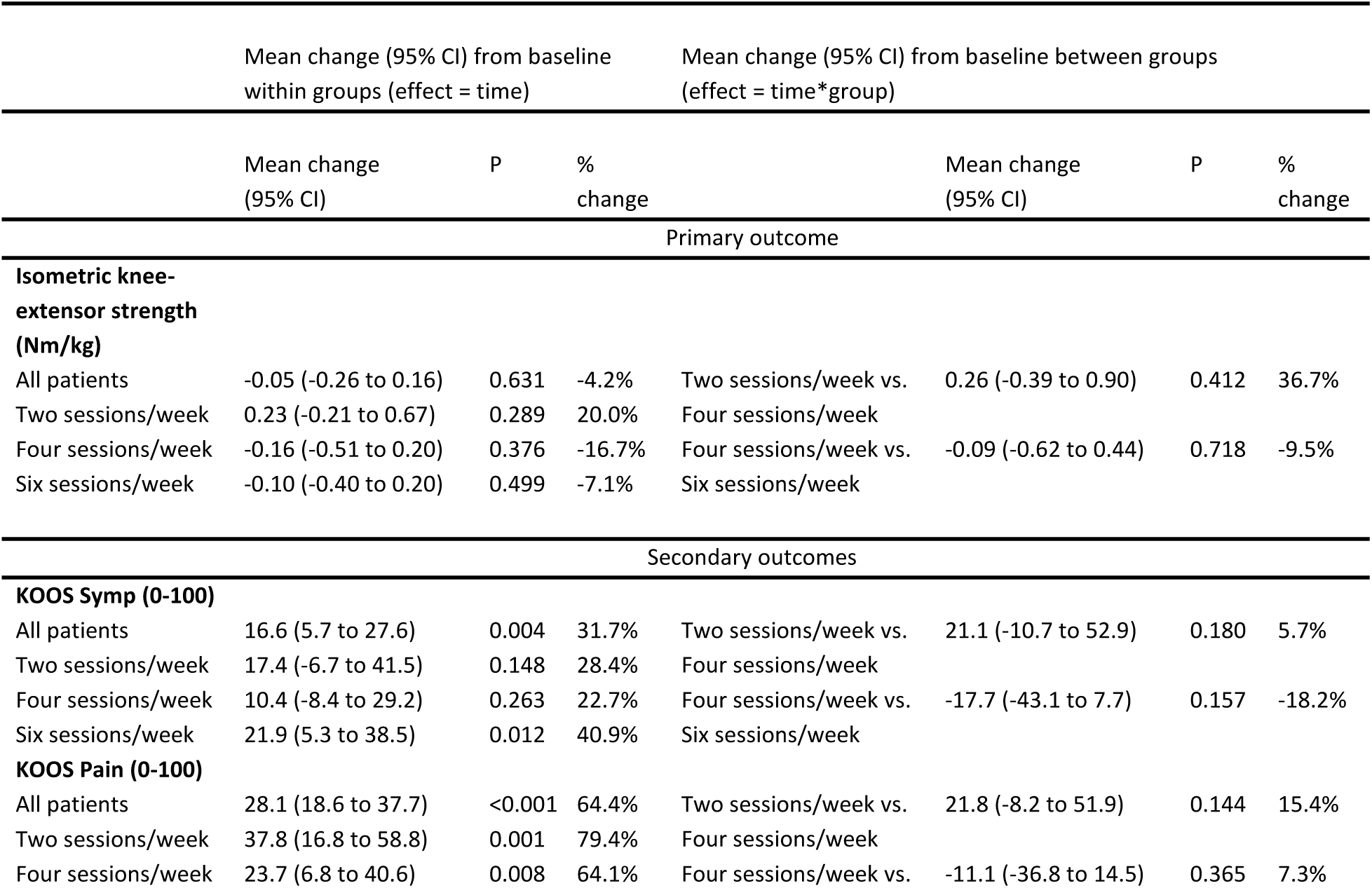
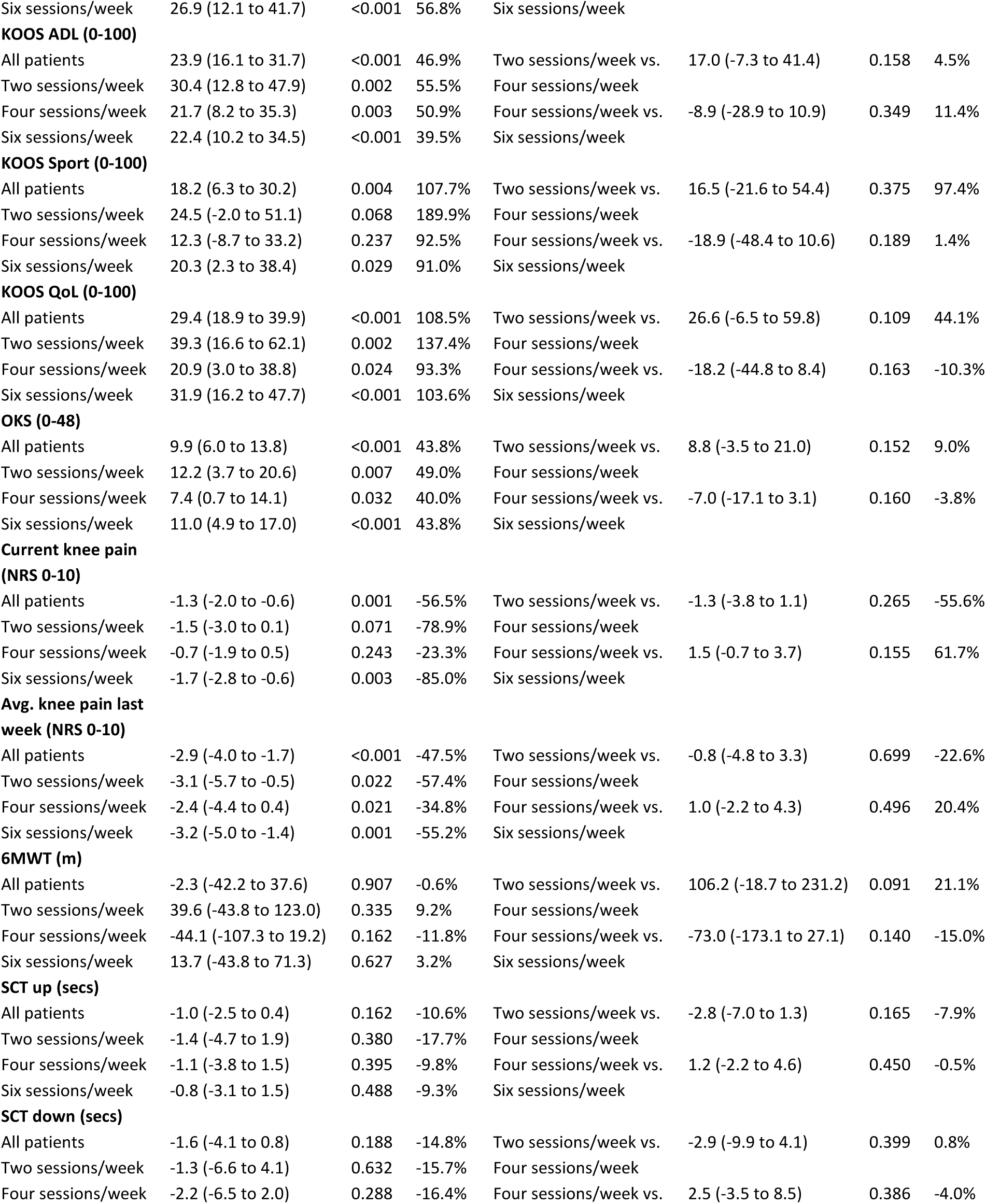
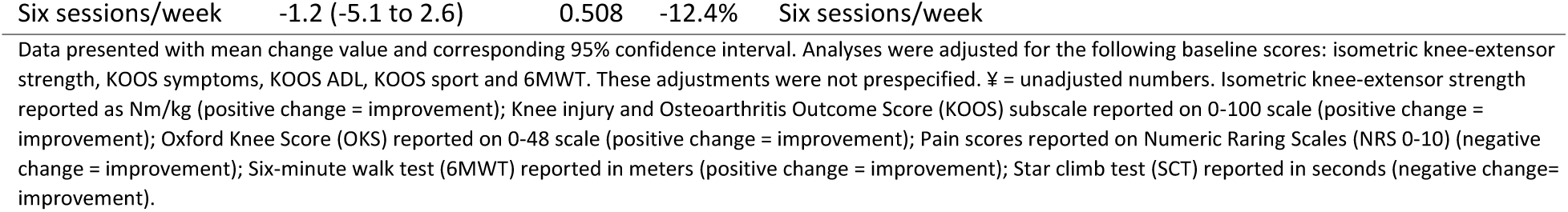
Mean change in all outcomes between baseline and three months after surgery (t_0_-t_3_). Intention-to-treat analysis, N = 32. One-way ANOVA based on imputed data.

**Secondary analyses (Mean change between group two and six sessions/week)**

**eTable 10.** Mean change between group two and six sessions/week for all outcomes between baseline and three months after surgery (t_0_-t_3_). Intention-to-treat analysis, N = 32. One-way ANOVA based on imputed data.

| Mean change (95% CI) from baseline between groups (effect = time*group) | Mean change (95% CI) | P | % change¥ |
| --- | --- | --- | --- |
| Two sessions/week vs. Six sessions/week |  |  |  |
| Primary outcome |  |  |  |
| **Isometric knee-extensor strength (Nm/kg)** | 0.12 (-0.46 to 0.69) | 0.681 | 27.1% |
| Secondary outcomes |  |  |  |
| **KOOS Symp (0-100)** | 1.9 (-27.1 to 30.9) | 0.889 | -12.4% |
| **KOOS Pain (0-100)** | 9.4 (-17.5 to 36.3) | 0.474 | 22.7% |
| **KOOS ADL (0-100)** | 7.1 (-14.7 to 28.9) | 0.503 | 16.0% |
| **KOOS Sport (0-100)** | -0.2 (-35.1 to 34.7) | 0.992 | 98.9% |
| **KOOS QoL (0-100)** | 7.3 (-22.9 to 37.6) | 0.620 | 33.8% |
| **OKS (0-48)** | 1.0 (-10.2 to 12.2) | 0.852 | 5.2% |
| **Current knee pain (NRS 0-10)** | -1.6 (-5.4 to 2.1) | 0.360 | 6.1% |
| **Avg. knee pain last week (NRS 0-10)** | 0.3 (-3.4 to 4.0) | 0.867 | -2.2% |
| **6MWT (m)** | 27.8 (-91.7 to 147.3) | 0.631 | 6.0% |
| **SCT up (secs)** | -1.7 (-5.5 to 2.1) | 0.371 | -8.4% |
| **SCT down (secs)** | -0.6 (-7.2 to 6.0) | 0.853 | -3.3% |

Data presented with mean change value and corresponding 95% confidence interval. Analyses were adjusted for the following baseline scores: isometric knee-extensor strength, KOOS symptoms, KOOS ADL, KOOS sport and 6MWT. These adjustments were not prespecified. ¥ = unadjusted numbers. Isometric knee-extensor strength reported as Nm/kg (positive change = improvement); Knee injury and Osteoarthritis Outcome Score (KOOS) subscale reported on 0-100 scale (positive change = improvement); Oxford Knee Score (OKS) reported on 0-48 scale (positive change = improvement); Pain scores reported on Numeric Rating Scales (NRS 0-10) (negative change = improvement); Six-minute walk test (6MWT) reported in meters (positive change = improvement); Star climb test (SCT) reported in seconds (negative change = improvement).

## Supplement 6. Co-morbidities disqualifying six patients for surgery after exercise

**eTable 11.** Co-morbidities disqualifying six patients for surgery after exercise (assessed by an orthopedic surgeon).

|  |  |
| --- | --- |
| 1 | Smoker, reduced lung function (diagnosed with chronic obstructive pulmonary disease) and obesity. |
| 2 | Acute neurological and urological diagnoses. |
| 3 | Obesity. |
| 4 | Acute neurological diagnosis. |
| 5 | Referred to further diagnosis. |
| 6 | Personal challenges. |

